# Device-assessed sleep and physical activity in individuals recovering from a hospital admission for COVID-19: a prospective, multicentre study

**DOI:** 10.1101/2022.02.03.22270391

**Authors:** the PHOSP-COVID Collaborative Group, Tatiana Plekhanova, Alex V Rowlands, Rachael A Evans, Charlotte L Edwardson, Nicolette C Bishop, Charlotte E Bolton, James D Chalmers, Melanie J Davies, Enya Daynes, Annemarie B Docherty, Omer Elneima, Neil J Greening, Sharlene A Greenwood, Andrew P Hall, Victoria C Harris, Ewen M Harrison, Joseph Henson, Ling-Pei Ho, Alex Horsley, Linzy Houchen-Wolloff, Kamlesh Khunti, Olivia C Leavy, Nazir I Lone, Michael Marks, Ben Maylor, Hamish J C McAuley, Claire M Nolan, Krisnah Poinasamy, Jennifer K Quint, Betty Raman, Matthew Richardson, Jack A Sargeant, Ruth M Saunders, Marco Sereno, Aarti Shikotra, Amisha Singapuri, Michael Steiner, David J Stensel, Louise V Wain, Julie Whitney, Dan G Wootton, Christopher E Brightling, William D-C Man, Sally J Singh, Tom Yates

## Abstract

**Objectives:** To describe physical behaviours following hospital admission for COVID-19 including associations with acute illness severity and ongoing symptoms.

**Methods:** 1077 patients with COVID-19 discharged from hospital between March and November 2020 were recruited. Using a 14-day wear protocol, wrist-worn accelerometers were sent to participants after a five-month follow-up assessment. Acute illness severity was assessed by the WHO clinical progression scale, and the severity of ongoing symptoms was assessed using four previously reported data-driven clinical recovery clusters. Two existing control populations of office workers and type 2 diabetes were comparators.

**Results:** Valid accelerometer data from 253 women and 462 men were included. Women engaged in a mean±SD of 14.9±14.7 minutes/day of moderate-to-vigorous physical activity (MVPA), with 725.6±104.9 minutes/day spent inactive and 7.22±1.08 hours/day asleep. The values for men were 21.0±22.3 and 755.5±102.8 minutes/day and 6.94±1.14 hours/day, respectively. Over 60% of women and men did not have any days containing a 30-minute bout of MVPA. Variability in sleep timing was approximately 2 hours in men and women. More severe acute illness was associated with lower total activity and MVPA in recovery. The very severe recovery cluster was associated with fewer days/week containing continuous bouts of MVPA, longer sleep duration, and higher variability in sleep timing. Patients post-hospitalisation with COVID-19 had lower levels of physical activity, greater sleep variability, and lower sleep efficiency than a similarly aged cohort of office workers or those with type 2 diabetes.

**Conclusions:** Physical activity and regulating sleep patterns are potential treatable traits for COVID-19 recovery programmes.

## INTRODUCTION

There have been over 330 million confirmed cases of COVID-19 and over 5.5 million deaths.^1^ Of the 15.6 million cases in the UK, there have been over 170,000 deaths and >660,000 patients admitted to hospital.^2^ As mortality improves, the number of post-hospitalisation survivors of COVID-19 is increasing. In some studies, more than 70% have not fully recovered by five months after discharge and have a substantial mental and physical health burden.^3^ Given this, the pressing need for research to inform and support rehabilitation post-hospitalisation with COVID-19 is evident.

Emerging evidence suggests that physical activity,^4,5^ good quality sleep, and regular sleep patterns^5^ are associated with lower odds of being admitted to hospital or dying with COVID-19. This may occur through a reduction in chronic inflammation^6,7^ and lower cardiometabolic risk factors, features associated with an increased risk of COVID-19,^8^ and/or through enhanced immunity.^9^ The ongoing burden of symptoms associated with poor recovery^3^ may have a detrimental impact on physical activity and sleep behaviours in post-hospitalisation survivors of COVID-19.

The post-hospitalisation COVID-19 (PHOSP-COVID) study is a large prospective multicentre follow-up study with the aim of understanding and improving long-term health outcomes following COVID-19 (https://phosp.org). Cluster analysis identified four recovery phenotypes relating to the severity of physical, mental, and cognitive health impairments an average of five months post-hospitalisation with COVID-19.^3^

The aim of this study was to describe accelerometer-assessed physical behaviours in patients post-hospitalisation with COVID-19 and to understand whether there are differences in physical behaviours associated with acute illness severity or the four recovery clusters. Physical behaviours within the PHOSP-COVID cohort were also described relative to a cohort of office workers and a cohort of adults with type 2 diabetes.

## METHODS

### Participants and methods

PHOSP-COVID is a prospective longitudinal cohort study recruiting patients aged ≥18 years who were discharged from 80 National Health Service (NHS) hospitals across England, Northern Ireland, Scotland, and Wales following admission to a medical assessment or ward for confirmed or clinician-diagnosed COVID-19. Participants were excluded if they: i) had a confirmed diagnosis of a pathogen unrelated to the objectives of this study, ii) attended an accident and emergency department but were not admitted, iii) had another life-limiting illness with life expectancy less than six months. This analysis included data from 1077 participants who attended a dedicated research visit at a median five-months (range 2-7 months) post-discharge between March and November 2020 previously described.^3^

All study participants provided written informed consent. The study was approved by the Leeds West Research Ethics Committee (20/YH/0225) and is registered on the ISRCTN Registry (ISRCTN10980107).

### Accelerometer data collection

Physical behaviours (i.e., physical activity and sleep) were assessed using the GENEActiv accelerometer (GENEActiv Original, ActivInsights, Kimbolton, UK). The monitors were initialised to record triaxial accelerations for 21 days at 30 Hz, with participants being asked to wear the monitor for 14 days.

Where possible, participants received the monitor and instructions by post within one month of their PHOSP-COVID research visit (2-7 months from discharge). Participants were instructed to start wearing the monitor on their non-dominant wrist immediately upon receiving it and to wear it 24 hours/day. Participants were asked to return their monitors in a prepaid envelope after the 14-day assessment period.

### Accelerometer data processing

Accelerometer files were processed with R-package GGIR version 2.2-0 (http://cran.r-project.org).^10^ Participants were excluded if they had <3 days of valid wear (defined as ≥16 hours/day).

Details of processing methods and definitions of the physical behaviour variables are shown in the *Supplementary material*.

Physical behaviour characteristics included average acceleration in m*g* (a proxy for physical activity volume), moderate-to-vigorous intensity physical activity (MVPA) accumulated in ≥1-minute bouts (minutes), light-intensity activity (minutes), inactive time (a proxy for sedentary time) in minutes, and intensity of the most active continuous 30 and 10 minutes/day (m*g*).

Weekly physical activity characteristics included the number of days/week with 10-and 30-minute continuous MVPA. Weekly variables were limited to participants with at least seven valid days of data.

Sleep characteristics included sleep duration (hours), sleep efficiency (%), and mid-sleep variability (within-person standard deviation of mid-sleep time). Sleep mid-point variability describes how variable people are in the timing of their sleep.

### Disease exposures

Acute illness severity was determined by the WHO clinical progression scale^11^ (*Supplementary material*).

The severity of ongoing symptoms after discharge was categorised on clusters derived previously within PHOSP-COVID where unsupervised machine learning using data from a battery of patient-reported outcomes and physical tests, identified four recovery clusters described by very severe, severe, moderate, and mild ongoing physical and mental health impairments.^3^ This outcome is referred to as recovery clusters.

Data for the severity of ongoing symptoms were missing in 27% of included participants due to missing data on the patient-reported outcomes and physical tests used for the cluster analysis.^3^

### Covariates

Data on sex, age at admission, ethnicity, number of chronic diseases, body mass index (BMI), and deprivation were included in this study (*Supplementary material*).

### Comparative Cohorts

In response to the lack of baseline data for the PHOSP-COVID cohort, the cohort was compared to accelerometer data collected in a cohort of office workers^12^ and individuals with type 2 diabetes^13,14^ for descriptive purposes. Details of the comparative cohorts are described in the *Supplementary material*. Accelerometer data from all three cohorts were processed using identical methods.

### Data selection

For this analysis, only those with valid accelerometer data were included. Of the 1077 participants in the PHOSP-COVID dataset, postal addresses were available to the research team for 853 participants within one month of follow-up clinical and research data collection visits allowing accelerometers to be posted, of which 796 were returned, with 715 providing valid data (*Supplementary Figure S1*).

### Statistical analysis

Differences in physical activity and sleep across acute illness severity and recovery clusters were assessed using generalised linear models. Continuous variables were analysed using a normal distribution with an identity link. Model selection was informed by the Akaike Information Criterion. Although some physical activity variables displayed non-parametric distributions, adjusted model fit was not meaningfully improved using different distribution or log links once covariates were added. Physical activity bout data was analysed using a Poisson distribution with a log-linear link as count data. Binary logistic regression was used to investigate the odds of not meeting 150 minutes of MVPA per week across acute illness severity and recovery clusters and reported as odds ratios (95% CI).

Data were adjusted for age at admission, sex, ethnicity, deprivation, number of comorbidities, season of data collection, number of wear days (activity outcomes) or wear nights (sleep outcomes). Interactions between sex and acute illness severity/recovery cluster were included to determine whether differences in physical activity or sleep variables across acute illness severity or recovery clusters varied by sex. Data are reported as sex-stratified marginal means (95% CI) derived from this model.

Generalised linear models were also used to examine associations between the variables that made up the ongoing severity cluster definitions (breathlessness, fatigue, anxiety, depression, post-traumatic stress disorder, physical performance, and cognition - *Supplementary material*) and physical activity and sleep characteristics. Variables were standardised and analysed as continuous variables. After generating the main effect for each exposure, cluster variable by sex interactions were added to the models and significant interactions were further stratified by sex. Data are reported as beta-coefficients (95% CI).

A sensitivity analysis was conducted, removing healthcare workers to examine whether healthcare work status had an impact on sleep variables.

Data were analysed using SPSS (version 26.0). A p-value of <0.05 was considered statistically significant.

## RESULTS

Of the 1077 participants included in the PHOSP-COVID dataset, 715 (253 women, 462 men) had valid accelerometer data. Participant characteristics are displayed in **Table 1**. 151 (32.7%) men and 54 (21.3%) women received invasive mechanical ventilation (IMV) during the acute illness (WHO class 7-9), and 172 (37.2%) men and 66 (26.1%) women were classified within the very severe recovery cluster. Participants’ characteristics with accelerometer data compared to those without are displayed in *Supplementary Table S1*. The proportion of the population within the different classifications and clusters of disease severity were similar in those with complete and missing accelerometer data. However, those with complete data were older (59 vs. 55 years), with a higher proportion from White ethnicities (69.8% vs. 58.3%) and the least deprived quintile (20.3% vs. 14.6%).

**Table 1:**
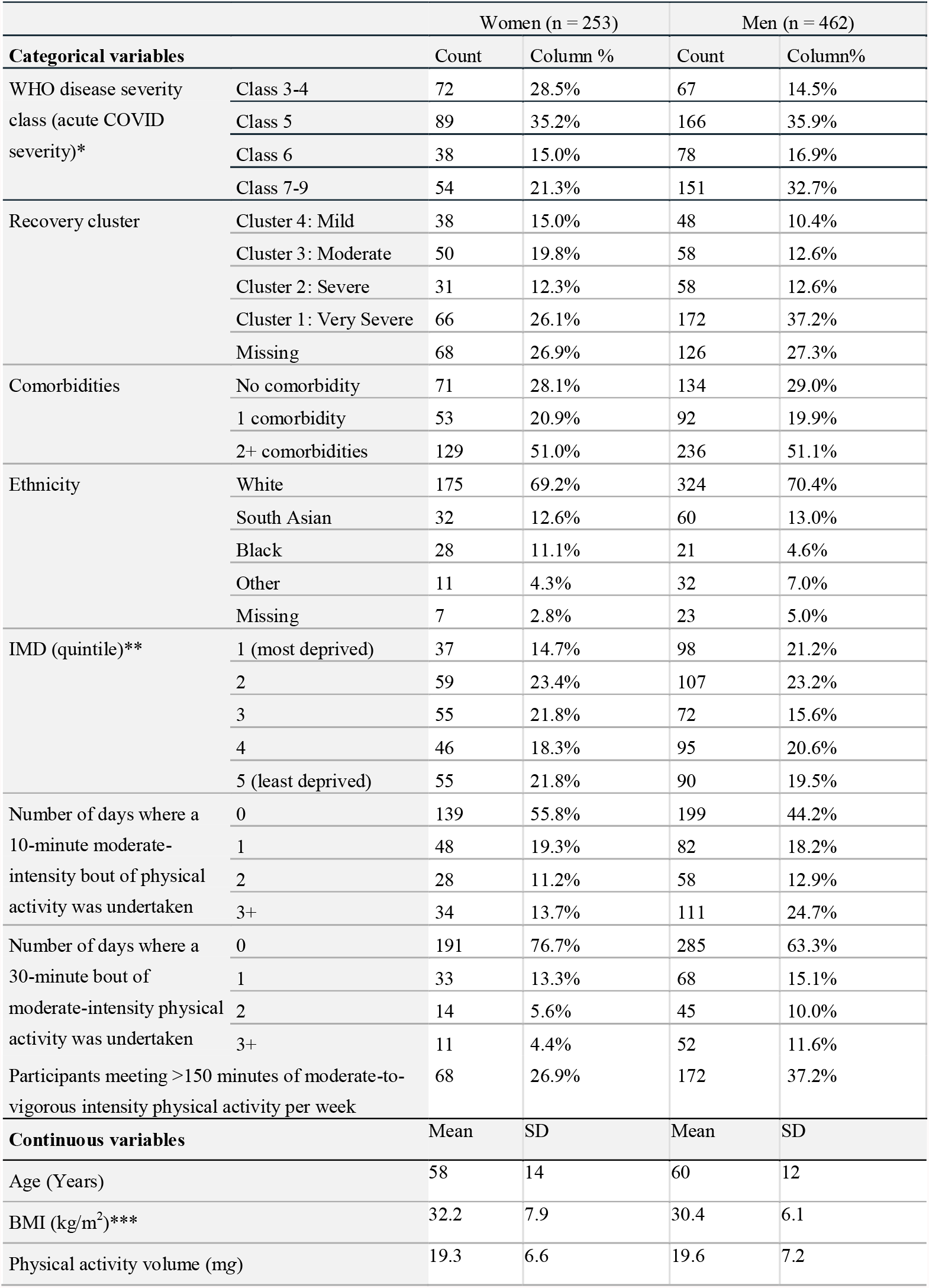
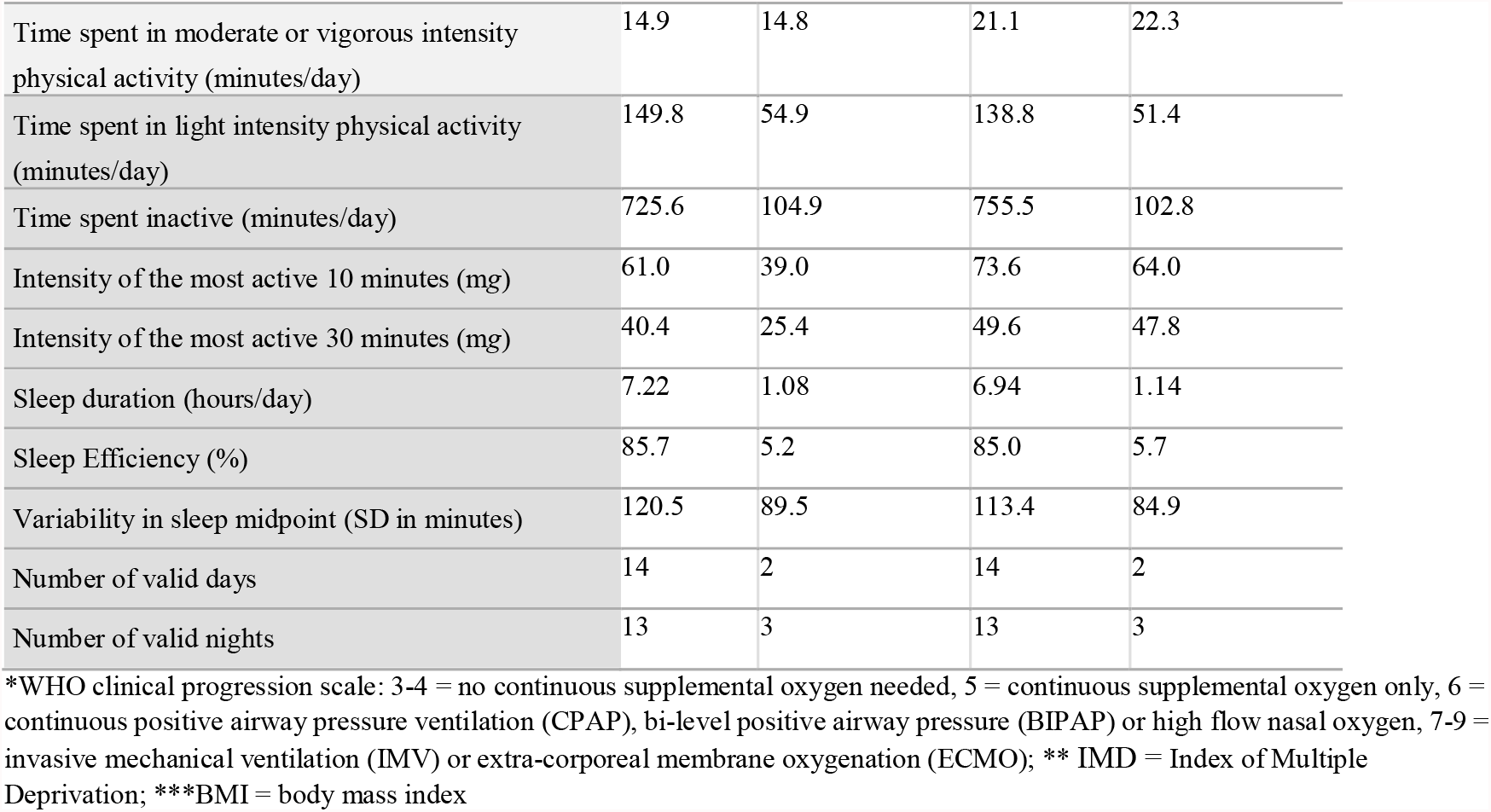
Participant Characteristics.

|  |  | Women (n = 253) |  | Men (n = 462) |  |
| --- | --- | --- | --- | --- | --- |
| **Categorical variables** |  | Count | Column % | Count | Column% |
| WHO disease severity class (acute COVID severity)* | Class 3-4 | 72 | 28.5% | 67 | 14.5% |
|  | Class 5 | 89 | 35.2% | 166 | 35.9% |
|  | Class 6 | 38 | 15.0% | 78 | 16.9% |
|  | Class 7-9 | 54 | 21.3% | 151 | 32.7% |
| Recovery cluster | Cluster 4: Mild | 38 | 15.0% | 48 | 10.4% |
|  | Cluster 3: Moderate | 50 | 19.8% | 58 | 12.6% |
|  | Cluster 2: Severe | 31 | 12.3% | 58 | 12.6% |
|  | Cluster 1: Very Severe | 66 | 26.1% | 172 | 37.2% |
|  | Missing | 68 | 26.9% | 126 | 27.3% |
| Comorbidities | No comorbidity | 71 | 28.1% | 134 | 29.0% |
|  | 1 comorbidity | 53 | 20.9% | 92 | 19.9% |
|  | 2+ comorbidities | 129 | 51.0% | 236 | 51.1% |
| Ethnicity | White | 175 | 69.2% | 324 | 70.4% |
|  | South Asian | 32 | 12.6% | 60 | 13.0% |
|  | Black | 28 | 11.1% | 21 | 4.6% |
|  | Other | 11 | 4.3% | 32 | 7.0% |
|  | Missing | 7 | 2.8% | 23 | 5.0% |
| IMD (quintile)** | 1 (most deprived) | 37 | 14.7% | 98 | 21.2% |
|  | 2 | 59 | 23.4% | 107 | 23.2% |
|  | 3 | 55 | 21.8% | 72 | 15.6% |
|  | 4 | 46 | 18.3% | 95 | 20.6% |
|  | 5 (least deprived) | 55 | 21.8% | 90 | 19.5% |
| Number of days where a 10-minute moderate-intensity bout of physical activity was undertaken | 0 | 139 | 55.8% | 199 | 44.2% |
|  | 1 | 48 | 19.3% | 82 | 18.2% |
|  | 2 | 28 | 11.2% | 58 | 12.9% |
|  | 3+ | 34 | 13.7% | 111 | 24.7% |
| Number of days where a 30-minute bout of moderate-intensity physical activity was undertaken | 0 | 191 | 76.7% | 285 | 63.3% |
|  | 1 | 33 | 13.3% | 68 | 15.1% |
|  | 2 | 14 | 5.6% | 45 | 10.0% |
|  | 3+ | 11 | 4.4% | 52 | 11.6% |
| Participants meeting >150 minutes of moderate-to-vigorous intensity physical activity per week |  | 68 | 26.9% | 172 | 37.2% |
| **Continuous variables** |  | Mean | SD | Mean | SD |
| Age (Years) |  | 58 | 14 | 60 | 12 |
| BMI (kg/m<sup>2</sup>)* |  | 32.2 | 7.9 | 30.4 | 6.1 |
| Physical activity volume (mg) |  | 19.3 | 6.6 | 19.6 | 7.2 |

|  |  |  |  |  |
| --- | --- | --- | --- | --- |
| Time spent in moderate or vigorous intensity physical activity (minutes/day) | 14.9 | 14.8 | 21.1 | 22.3 |
| Time spent in light intensity physical activity (minutes/day) | 149.8 | 54.9 | 138.8 | 51.4 |
| Time spent inactive (minutes/day) | 725.6 | 104.9 | 755.5 | 102.8 |
| Intensity of the most active 10 minutes (mg) | 61.0 | 39.0 | 73.6 | 64.0 |
| Intensity of the most active 30 minutes (mg) | 40.4 | 25.4 | 49.6 | 47.8 |
| Sleep duration (hours/day) | 7.22 | 1.08 | 6.94 | 1.14 |
| Sleep Efficiency (%) | 85.7 | 5.2 | 85.0 | 5.7 |
| Variability in sleep midpoint (SD in minutes) | 120.5 | 89.5 | 113.4 | 84.9 |
| Number of valid days | 14 | 2 | 14 | 2 |
| Number of valid nights | 13 | 3 | 13 | 3 |

\*WHO clinical progression scale: 3-4 = no continuous supplemental oxygen needed, 5 = continuous supplemental oxygen only, 6 = continuous positive airway pressure ventilation (CPAP), bi-level positive airway pressure (BIPAP) or high flow nasal oxygen, 7-9 = invasive mechanical ventilation (IMV) or extra-corporeal membrane oxygenation (ECMO); \*\* IMD = Index of Multiple Deprivation; \*\*\*BMI = body mass index

The summary variables from the accelerometer data for women and men are displayed in **Table 1**. The median time from discharge to accelerometer wear was 245 days [IQR 178–276 days]. The median time from the PHOSP-COVID research visit to accelerometer wear was 65 days [IQR 11–93 days]. Accelerometer data were available for a mean of 14 valid days. Women engaged in a mean±SD of 14.9±14.7 minutes/day of MVPA, with 725.6±104.9 minutes/day spent inactive and 7.22±1.08 hours/day asleep. The same values for men were 21.0±22.3 and 755.5±102.8 minutes/day and 6.94±1.14 hours/day, respectively. Variability in sleep midpoint was ∼2 hours in men and women. Over 60% of both women and men did not have any days in a week that contained a 30-minute bout of MVPA, e.g., walking, with most women (56%) also not having any days with a bout of 10-minute of MVPA **(Table 1)**.

### Associations with disease severity

Across acute illness severity, those who had the most severe acute illness had ∼1-2 m*g* lower volume of physical activity (p=0.045) and less time spent in MVPA (p=0.032) (**Table 2**). Women who received IMV undertook the lowest levels of MVPA [13.7 minutes/day; 95% CI 7.3, 20.2] (**Table 2**). Women and men with the most severe acute disease were 3.38 (95% CI 1.29, 8.85) and 2.17 (95% CI 1.06, 4.45) times more likely, respectively, to not meet physical activity recommendations for health compared to those with the least severe disease (*Supplementary Figure S2*). The pattern of number of days/week with continuous bouts of MVPA was similar across acute illness severity (**Figure 1**).

**Table 2:** Physical activity and sleep variables across WHO classes of acute illness severity.

|  | Women |  |  |  | Men |  |  |  |  |  |
| --- | --- | --- | --- | --- | --- | --- | --- | --- | --- | --- |
| Physical activity variables | Class 3-4 | Class 5 | Class 6 | Class 7-9 | Class 3-4 | Class 5 | Class 6 | Class 7-9 | P for class | P for class x sex |
| Physical activity volume (mg) | 21.29 (19.26, 23.32) | 22.33 (20.43, 24.23) | 19.91 (17.4, 22.42) | 19.81 (17.58, 22.04) | 21.91 (19.82, 24) | 22.59 (20.98, 24.2) | 23.31 (21.24, 25.39) | 21.04 (19.29, 22.79) | 0.045 | 0.248 |
| Moderate to vigorous intensity physical activity (minutes/day) | 18.71 (12.86, 24.56) | 19.02 (13.55, 24.5) | 15.95 (8.72, 23.18) | 13.74 (7.31, 20.17) | 25.08 (19.06, 31.11) | 28.50 (23.85, 33.15) | 27.04 (21.06, 33.02) | 21.61 (16.56, 26.66) | 0.032 | 0.783 |
| Light intensity physical activity (minutes/day) | 167.76 (152.03, 183.49) | 170.95 (156.22, 185.67) | 150.16 (130.72, 169.6) | 158.2 (140.69, 175.72) | 153.99 (137.79, 170.19) | 153.77 (141.26, 166.28) | 160.73 (144.59, 176.88) | 149.12 (135.46, 162.78) | 0.386 | 0.150 |
| Inactivity (minutes/day) | 722.24 (692.72, 751.76) | 734.84 (707.2, 762.47) | 739.77 (703.29, 776.25) | 749.94 (717.08, 782.81) | 764.91 (734.5, 795.32) | 752.39 (728.92, 775.87) | 751.2 (720.9, 781.5) | 777.88 (752.24, 803.51) | 0.201 | 0.569 |
| Intensity of the most active continuous 30 minutes (mg) | 54.60 (42.06, 67.15) | 55.85 (44.19, 67.5) | 47.77 (32.23, 63.31) | 51.65 (37.97, 65.33) | 62.83 (49.98, 75.68) | 66.51 (56.53, 76.48) | 62.94 (50.16, 75.71) | 63.05 (52.16, 73.94) | 0.659 | 0.934 |
| Intensity of the most active continuous 10 minutes (mg) | 76.75 (59.59, 93.91) | 85.28 (69.33, 101.22) | 71.35 (50.09, 92.61) | 74.74 (56.03, 93.45) | 94.67 (77.09, 112.25) | 98.82 (85.17, 112.47) | 96.61 (79.13, 114.09) | 88.47 (73.57, 103.36) | 0.334 | 0.831 |
| **Sleep variables** |  |  |  |  |  |  |  |  |  |  |
| Sleep duration (hrs/night) | 6.95 (6.62, 7.29) | 6.84 (6.53, 7.15) | 7.02 (6.61, 7.42) | 6.72 (6.35, 7.08) | 6.58 (6.24, 6.92) | 6.54 (6.27, 6.8) | 6.48 (6.15, 6.82) | 6.46 (6.18, 6.75) | 0.514 | 0.771 |
| Sleep efficiency (%) | 86.75 (85.08, 88.42) | 86.12 (84.56, 87.68) | 85.89 (83.83, 87.95) | 85.78 (83.95, 87.61) | 85.94 (84.23, 87.66) | 85.35 (84.02, 86.67) | 84.78 (83.07, 86.48) | 85.46 (84.01, 86.9) | 0.524 | 0.951 |
| Sleep midpoint variability (minutes) | 91.8 (66.05, 117.55) | 92.72 (68.66, 116.78) | 88.19 (56.44, 119.95) | 113.65 (85.4, 141.91) | 95.55 (69.07, 122.02) | 91.9 (71.48, 112.31) | 106.88 (80.58, 133.18) | 85.7 (63.46, 107.95) | 0.854 | 0.151 |

Data reported as marginal mean (95% CI). Adjusted for age, sex, ethnicity, deprivation, number of comorbidities, season of data collection and number of wear days (physical activity variables) or wear nights (sleep variables). Models included 675 participants with complete cluster and covariate data

**Figure 1:**
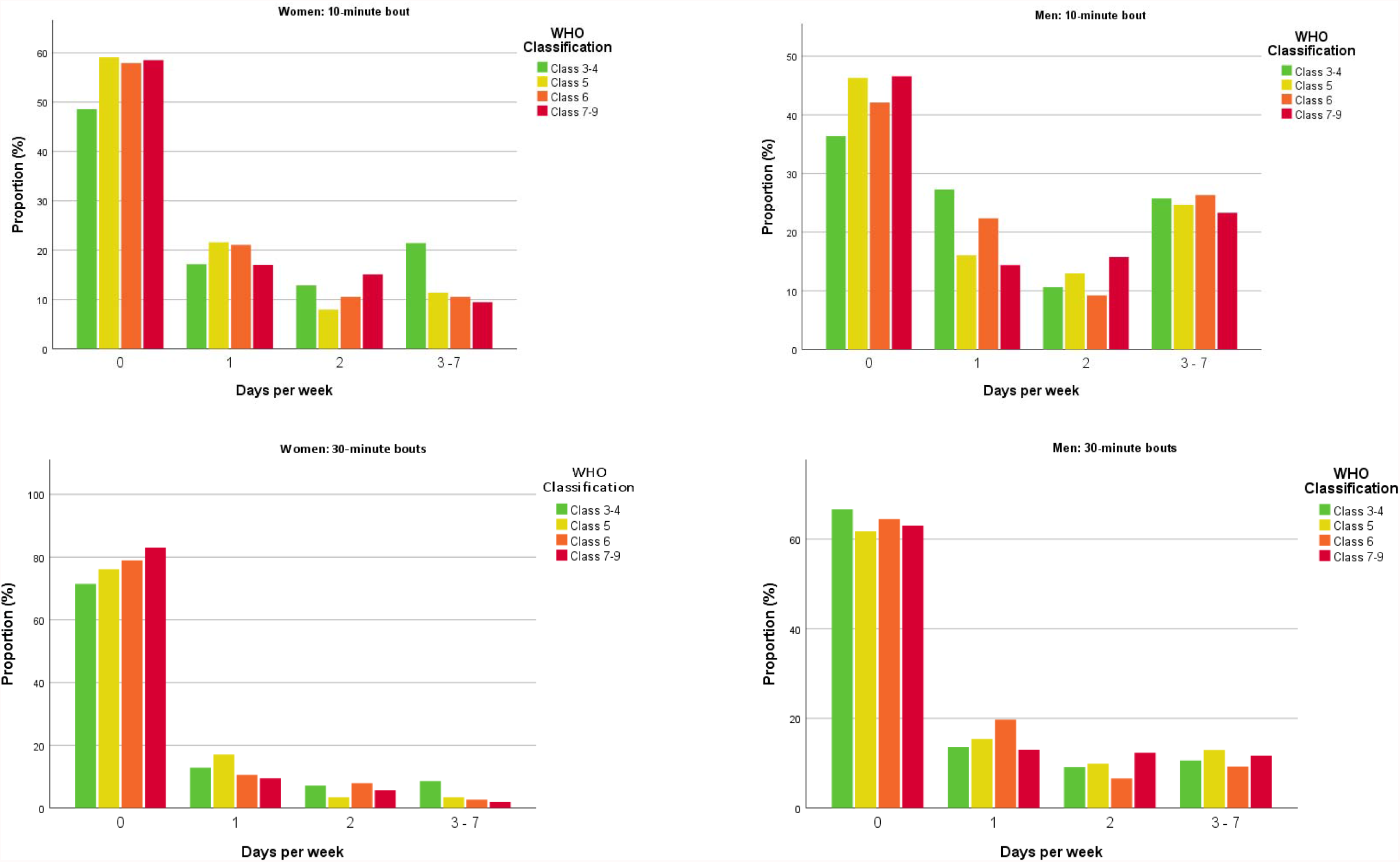
Proportion of participants within each WHO class of COVID-19 severity undertaking continuous bouts of physical activity Data display the proportion within each WHO class achieving 0, 1, 2 and 3-7 days per week with a bout of 10 minutes (top panel) or 30 minutes (bottom) at least moderate-intensity physical activity. Adjusted for age, sex, ethnicity, deprivation, number of comorbidities, season of data collection, and number of wear days. <u>10-minute bouts</u>: p for difference by cluster = 0.132, p for sex x cluster = 0.395. <u>30-minute bouts:</u> p for difference by cluster = 0.217, p for sex x cluster = 0.128

Across recovery clusters, there was no difference in daily volume of physical activity and time spent in light activity or MVPA (**Table 3**). However, time spent inactive was greater in men than women (p=0.013), with men in the very severe recovery cluster spending the most time inactive [789.05 minutes/day; 95% CI 753.78, 824.32]. Men in the severe and the very severe recovery clusters were also 2.52 (95% CI 1.19, 5.36) and 3.48 (95% CI 1.41, 8.59) times more likely, respectively, to not meet physical activity recommendations for health compared to those in the less severe recovery clusters (*Supplementary Figure S3*).

**Table 3:** Physical activity and sleep variables across four recovery clusters.

|  | Women |  |  |  | Men |  |  |  |  |  |
| --- | --- | --- | --- | --- | --- | --- | --- | --- | --- | --- |
| Physical activity variables | Cluster 4: Mild | Cluster 3: Moderate | Cluster 2: Severe | Cluster 1: Very Severe | Cluster 4: Mild | Cluster 3: Moderate | Cluster 2: Severe | Cluster 1: Very Severe | P for class | P for class x sex |
| Physical activity volume (mg) | 21.54 (19.49, 23.59) | 22.19 (19.52, 24.86) | 19.34 (16.93, 21.75) | 20.61 (18.12, 23.1) | 22.35 (20.64, 24.07) | 22.34 (20.22, 24.46) | 22.72 (20.52, 24.93) | 21.08 (18.63, 23.52) | 0.395 | 0.293 |
| Moderate to vigorous intensity physical activity (minutes/day) | 19.21 (13.02, 25.39) | 18.33 (10.26, 26.4) | 16.03 (8.75, 23.31) | 16.76 (9.24, 24.27) | 29.31 (24.13, 34.5) | 26.74 (20.34, 33.14) | 23.97 (17.31, 30.63) | 21.22 (13.84, 28.59) | 0.156 | 0.756 |
| Light intensity physical activity (minutes/day) | 164.84 (149.6, 180.07) | 168.15 (148.41, 187.89) | 145.21 (127.39, 163.02) | 156.29 (137.88, 174.69) | 148.08 (135.38, 160.77) | 153.17 (137.49, 168.85) | 158.33 (141.86, 174.81) | 139.25 (121.19, 157.31) | 0.357 | 0.065 |
| Inactivity (minutes/day) | 730.23 (700.48, 759.99) | 714.69 (676.13, 753.26) | 751.98 (717.18, 786.79) | 747.73 (711.78, 783.68) | 767.41 (742.61, 792.2) | 765.02 (734.39, 795.64) | 723.71 (691.53, 755.89) | 789.05 (753.78, 824.32) | 0.131 | 0.013 |
| Intensity of the most active continuous 30 minutes (mg) | 56.18 (41.87, 70.48) | 59.61 (41.15, 78.07) | 53.02 (36.28, 69.75) | 45.19 (27.93, 62.44) | 70.42 (58.43, 82.42) | 69.29 (54.5, 84.08) | 69.44 (53.84, 85.04) | 55.91 (38.64, 73.18) | 0.170 | 0.954 |
| Intensity of the most active continuous 10 minutes (mg) | 86.1 (66.6, 105.61) | 84.55 (59.38, 109.72) | 78.16 (55.33, 100.98) | 67.03 (43.50, 90.55) | 104.6 (88.25, 120.96) | 97.43 (77.26, 117.6) | 95.58 (74.31, 116.86) | 80.51 (56.96, 104.05) | 0.079 | 0.982 |
| **Sleep variables** |  |  |  |  |  |  |  |  |  |  |
| Sleep duration (hrs/night) | 6.97 (6.64, 7.3) | 6.8 (6.36, 7.23) | 7.20 (6.81, 7.59) | 6.85 (6.44, 7.25) | 6.52 (6.24, 6.8) | 6.41 (6.07, 6.76) | 6.86 (6.5, 7.23) | 6.52 (6.12, 6.92) | 0.039 | 0.960 |
| Sleep efficiency (%) | 85.86 (84.15, 87.56) | 85.08 (82.85, 87.31) | 87.25 (85.24, 89.26) | 86.65 (84.58, 88.72) | 85.62 (84.19, 87.05) | 84.58 (82.81, 86.34) | 85.23 (83.37, 87.08) | 85.14 (83.1, 87.17) | 0.412 | 0.534 |
| Sleep midpoint variability (minutes) | 92.44 (66.31, 87.56) | 95.29 (61.11, 87.31) | 74.46 (43.69, 89.26) | 112.02 (80.26, 88.72) | 87.44 (65.55, 87.05) | 97.83 (70.74, 86.34) | 78.85 (50.41, 87.08) | 105.53 (74.28, 87.17) | 0.070 | 0.956 |

Data reported as marginal mean (95% CI). Adjusted for age, sex, ethnicity, deprivation, number of comorbidities, season of data collection and number of wear days (physical activity variables) or wear nights (sleep variables). Models included 490 participants with complete cluster and covariate data

There was also a notable difference in the number of days/week on which longer bouts of physical activity were undertaken. In the very severe recovery cluster, over 80% of women and men did not undertake a bout of MVPA lasting 30 minutes on any day of the week, with over 60% not undertaking a bout lasting 10 minutes on any day (**Figure 2**). The frequency of longer bouts of MVPA was substantially higher in those within the mild recovery cluster; over 20% in the mild cluster undertook at least a 10-minute bout of MVPA on at least 3 days/week (**Figure 2**). Men and women in the severe recovery cluster had the longest sleep duration (p=0.039), with the very severe recovery cluster having the greatest sleep midpoint variability, although this did not reach significance (p=0.070) (**Table 3**).

**Figure 2:**
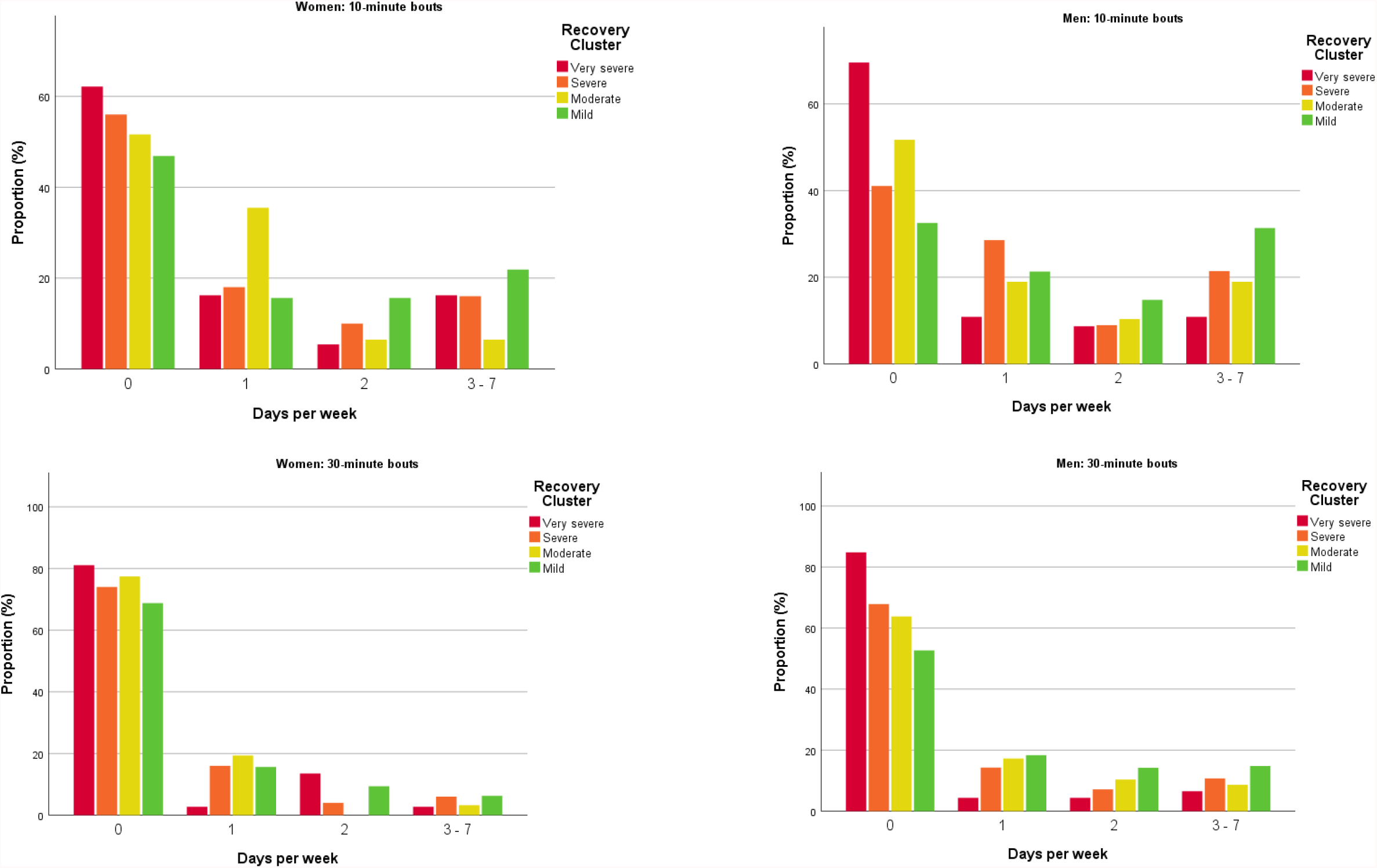
Proportion of participants within each recovery cluster undertaking continuous bouts of physical activity. Data display the proportion within each cluster achieving 0, 1, 2 and 3-7 days per week with a bout of 10 minutes (top panel) or 30 minutes (bottom) at least moderate-intensity physical activity. Adjusted for age, sex, ethnicity, deprivation, number of comorbidities, season of data collection, and number of wear days. <u>10-minute bouts</u>: p for difference by cluster < 0.001, p for sex x cluster = 0.672. <u>30-minute bouts</u>: p for difference by cluster = 0.021, p for sex x cluster = 0.262

Sleep variables across acute illness severity and recovery clusters showed a similar pattern when removing healthcare workers (N=98) (*Supplementary Table S2*).

The associations of recovery cluster variables with physical activity and sleep characteristics are shown in **Figure 3** (data shown in *Supplementary Table S3*). Lower severity of symptoms, except for cognition and anxiety, were positively associated with physical activity (p<0.05). More severe breathlessness, fatigue, anxiety, depression, and post-traumatic stress disorder were all associated with greater sleep midpoint variability (p<0.01). More severe depression was also associated with lower sleep efficiency (p=0.039). Associations of physical performance with MVPA and intensity of the most active continuous 30/10 minutes were stronger in men than women (p for interaction <0.05) (*Supplementary Table S3 and S4*).

**Figure 3:**
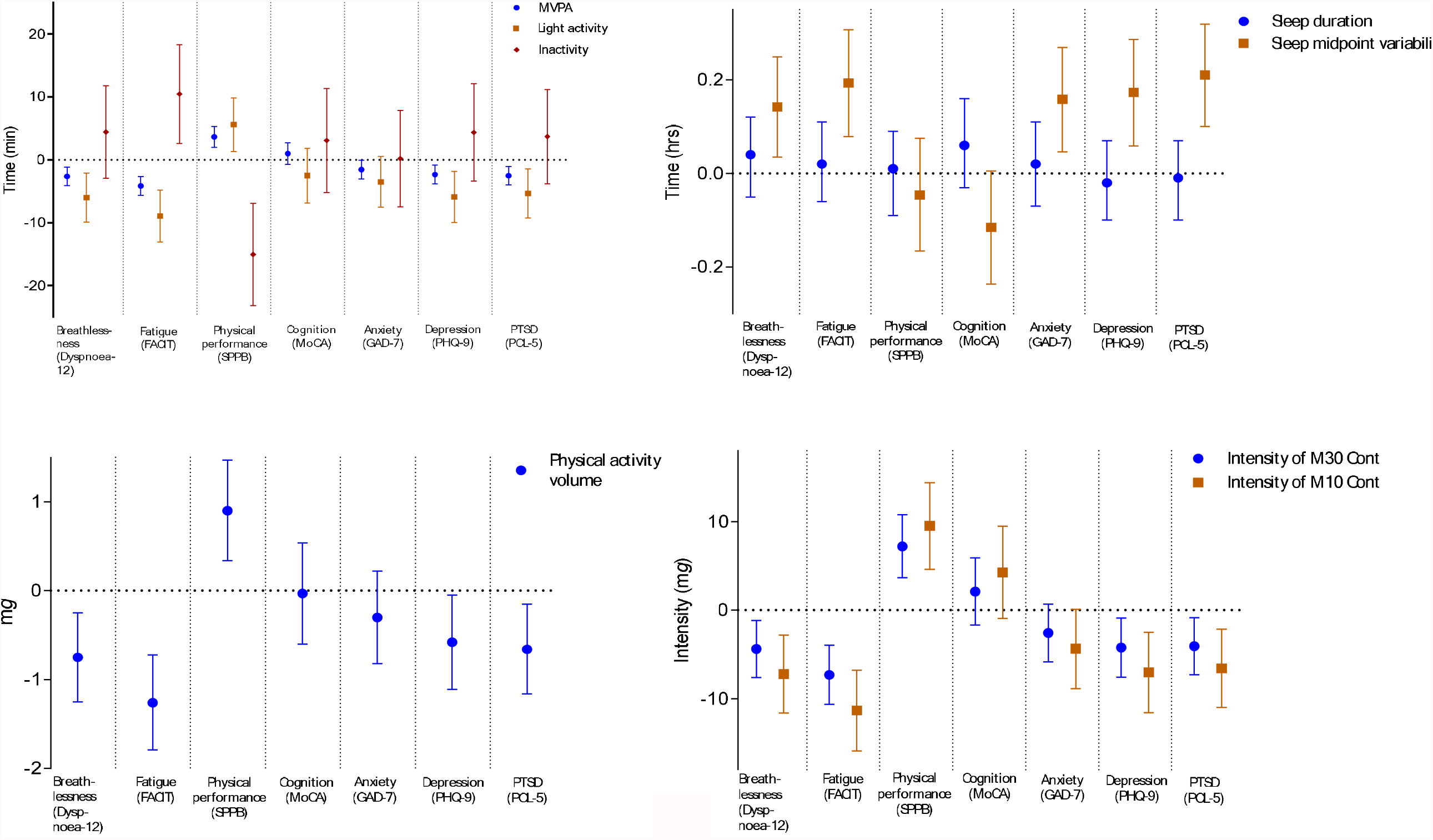
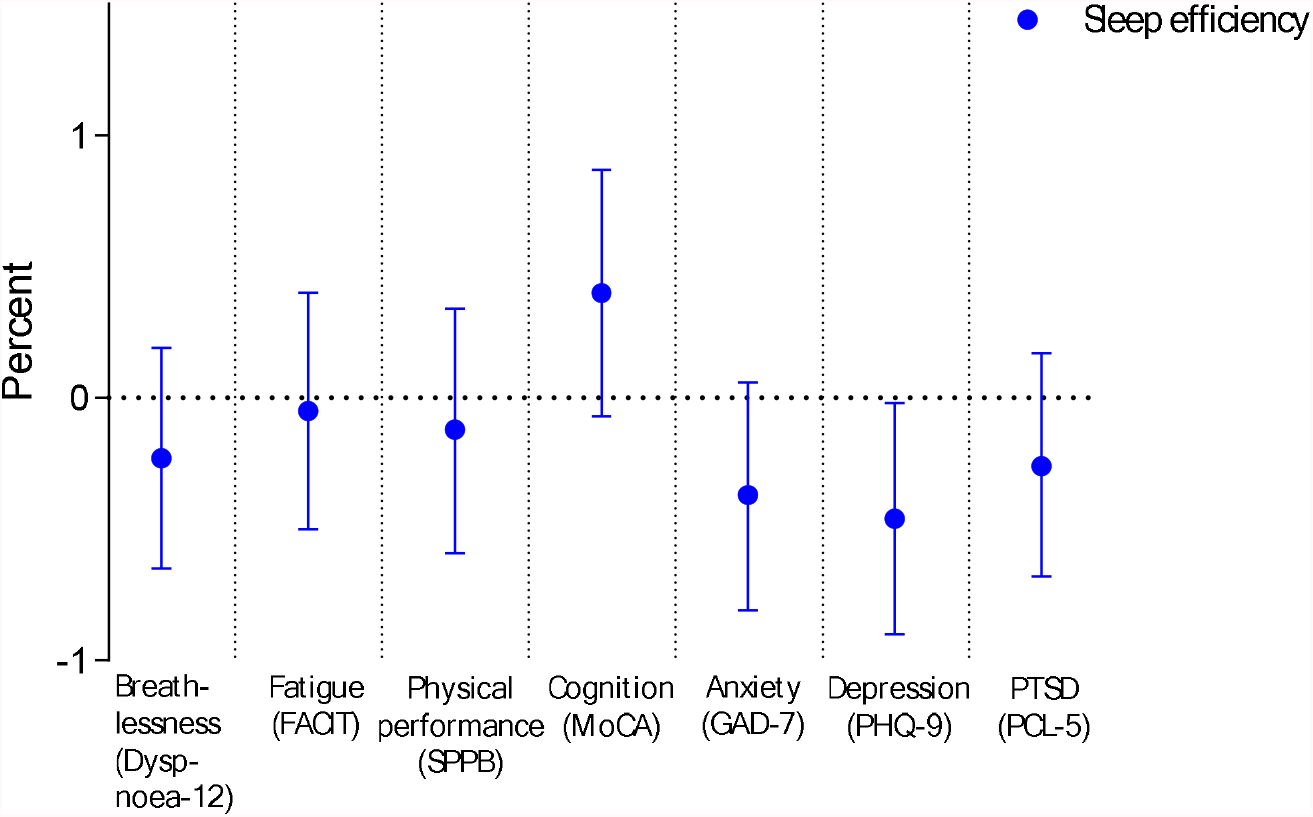
Forest plots of associations between patient-reported outcomes with physical activity and sleep characteristics. Data are shown as beta-coefficients (95% CI) showing the difference per SD in the exposure. Adjusted for age, sex, ethnicity, deprivation, number of comorbidities, season of data collection, and number of wear days (physical activity variables) or nights (sleep variables). FACIT = Functional Assessment of Chronic Illness Therapy; SPPB = short physical performance battery; MoCA = Montreal Cognitive Assessment; GAD-7 = General Anxiety Disorder 7 Questionnaire; PHQ-9 = Patient Health Questionnaire – 9; PTSD = post-traumatic stress disorder; PCL-5 = Post Traumatic Stress Disorder Checklist.

### Comparison to cohorts of office workers and those with type 2 diabetes

Characteristics of the SMART Work and Life (SWL) and Chronotype of Patients with Type 2 Diabetes and Effect on Glycaemic Control (CODEC) comparator cohorts are shown in *Supplementary Table S5* with differences in physical behaviours shown in *Supplementary Table S6*. Mean age was within five years of the PHOSP-COVID cohort for both comparator cohorts, while the CODEC cohort was well-matched for key characteristics including sex, multimorbidity status and BMI. Overall, activity was higher in the SWL and CODEC cohorts compared to PHOSP-COVID, with notably higher activity in the SWL cohort. The differences in activity volume of 1.1 m*g* in CODEC and 3.2 m*g* in SWL. Those in PHOSP-COVID spent ∼17 fewer minutes in light-intensity activity than those in CODEC (p=0.004) and ∼16 fewer minutes in MVPA (p<0.001) than those in SWL, with men also spending more time inactive compared to CODEC (p=0.020). The intensity of the most active 30/10 minutes was also lowest in PHOSP-COVID (p<0.001). The frequency of continuous bouts of MVPA/week in PHOSP-COVID was similar to CODEC, but notably lower compared to SWL (p<0.001) (*Supplementary Figure S4*). The variability in sleep midpoint was at least three times greater in PHOSP-COVID compared to SWL and CODEC (p<0.001), with sleep efficiency also lower (p<0.001) being 83.5% [95%CI 82.3, 84.7] in women and 82.7% [95%CI 81.6, 83.9] in men, but ≥86.5% in SWL and CODEC. Sleep duration was similar across the three cohorts (p>0.092).

## DISCUSSION

Women and men recovering from a hospital admission for COVID-19 had low levels of physical activity and high variability in sleep timing. More severe acute illness was associated with a lower volume of physical activity (approximating 500-1000 fewer steps per day)^15^ and fewer minutes accumulated in MVPA, whereas more severe recovery clusters were associated with a low frequency of continuous sessions of physical activity per week, longer sleep duration and high variability in sleep timing. Further, the physical activity and sleep profile were notably worse in the PHOSP-COVID cohort than a similarly aged office worker comparator cohort,^12^ with the 3.2 m*g* difference in activity volume approximating 1600 fewer steps per day.^15^ Relative to our well-matched comparator group with type 2 diabetes,^14^ differences in activity volume approximated 550 fewer steps per day,^15^ approximately 17 fewer minutes spent in light-intensity activity, with three times higher variability in sleep timing and lower sleep efficiency.

Among COVID-19 sufferers, sleep disturbance is one of the most commonly reported symptoms, irrespective of acute illness severity, and is highly prevalent following hospital discharge.^16^ Irregular sleep patterns as observed in this cohort, independent of sleep duration, can lead to circadian disruption which is a risk factor for metabolic syndrome, obesity, dyslipidemia, and diabetes.^17^

Being inactive, defined as not meeting the physical activity guidelines of 150 minutes of MVPA per week,^18^ is a risk factor for acute COVID-19 severity, with those who are inactive being 2.26 times more likely to be admitted to hospital, 1.73 times more likely to need intensive care, and 2.49 times more likely to die.^4^ Given this, and that habitual physical activity is generally fairly stable,^19^ it is possible that the lower physical activity in those with more severe acute COVID-19 may reflect their activity levels prior to infection with COVID-19.

The recovery clusters in PHOSP-COVID are not closely associated with acute illness severity,^3^ consistent with previous research.^20^ Thus, the differences observed in continuous bouts of physical activity and sleep across recovery clusters likely reflect the participants’ current mental and physical health impairment. This suggests that the ability to sustain a 10-or 30-minute session of activity without resting is compromised in those with more severe ongoing symptoms. Further, the sleep routine appears to be disrupted across all participants within the dataset, but particularly in those in the more severe cluster. Both behaviours are associated with the multiple impairments that characterise the more severe clusters,^3^ including anxiety and depression,^21-23^ fatigue,^24^ and physical function.^25^ These data suggest that rehabilitation pathways that have been set up to manage recovery from COVID-19 should focus on the spectrum of behaviours that encompass the 24-hour period, including facilitating a return to normal patterns of physical activity, including being able to undertake longer bouts of physical activity, along with focusing on addressing sleep disruption issues.

### Strengths and limitations

Key strengths of this study are its size, the comprehensively phenotyped multicentre cohort with novel clinical phenotypes, and accelerometer-assessed physical behaviours at scale. Irrespective, the study has several limitations. Notably, it was not possible to obtain measures of physical behaviours for the participants before they were infected with COVID-19. To account for this, we compared the data to a similarly aged cohort of office workers and a cohort of adults with type 2 diabetes who were well-matched on key characteristics including sex, multimorbidity status, and BMI.^13,14^ However, the data were collected on the comparator cohorts prior to the pandemic; we acknowledge that patterns of physical behaviours may also have been impacted due to the COVID-19 restrictions that have been imposed in the UK (and worldwide) to limit the spread of the virus. Variability in sleep timing and sleep efficiency were the main differences between the PHOSP-COVID and comparator cohorts in the present study. We have previously shown that these sleep-related variables did not differ before and during COVID-19 restrictions, suggesting the differences observed are unlikely due to differences in the measurement period.^26^ Due to missing patient-reported outcome data within the PHOSP-COVID cohort, a cluster assignment was not derived for all participants. Finally, over 66% of the cohort had valid accelerometer data but those with data tended to be older, from less deprived communities, and with a lower proportion from ethnic minority communities. Therefore, the data presented may not be generalizable to all those recovering from a hospital admission for COVID-19. Although we used data from a well-phenotyped cohort of patients recovering from a hospital admission for COVID-19, it is not possible to disentangle to what extent these results are specific to COVID-19 or reflect recovery from acute illness requiring hospitalisation more generally.

## Conclusion

Survivors of a hospital admission for COVID-19 have low levels of physical activity and significantly disrupted patterns of sleep several months after discharge. Acute illness severity was associated with lower total and moderate-to-vigorous activity following discharge, whereas more severe recovery clusters were associated with substantially fewer bouts of continuous physical activity and greater variability in sleep timing. Without modification, these behaviours are likely to result in further future disease. Physical activity, particularly sustained continuous bouts, and variability in sleep timing are potential treatable traits for survivors of COVID-19.

### What are the new findings?

- Survivors of COVID-19 had low levels of physical activity and disrupted patterns of sleep eight months post-discharge.
- More severe acute illness was associated with a lower volume of physical activity and fewer minutes accumulated in moderate-to-vigorous intensity physical activity
- More severe ongoing symptoms were associated with a low frequency of continuous bouts of physical activity per week and high variability in sleep timing.
- Survivors of COVID-19 had lower levels of physical activity and a worse sleep profile compared to a similarly aged cohort of office workers or those with type 2 diabetes.

### How might it impact on clinical practice in the future?

- Our findings highlight the potential importance of addressing sleep patterns and the ability to sustain continuous movement in future interventions in those recovering from severe COVID-19.

## Supporting information

PHOSP-COVID Collaborative Group

## Data Availability

The protocol, consent form, definition and derivation of clinical characteristics and outcomes, training materials, regulatory documents, information about requests for data access, and other relevant study materials are available online https://www.phosp.org/.

## Ethics statements

### Patient consent for publication

Not required.

### Ethics approval

The study was approved by the Leeds West Research Ethics Committee (20/YH/0225).

## Author contributions

The manuscript was initially drafted by TP, TY, AVR, and further developed by the writing committee. TY, AVR, CLE, TP made substantial contributions to the conception and design of the work. TP, CLE, AVR, RAE, ASi, MS, RMS, VCH, CEBo, JDC, DGW, L-PH, AH, MM, WD-CM made substantial contributions to the acquisition of data. TY, AVR, TP, CLE, RAE, MJD made contributions to the analysis or interpretation of data for the work. TY, AVR, TP verified the underlying data. All authors contributed to data interpretation and critical review and revision of the manuscript. All authors had full access to all the data in the study and had final responsibility for the decision to submit for publication.

## Acknowledgements

This study would not be possible without all the participants who have given their time and support. We thank all the participants and their families. We thank the many research administrators, health-care and social-care professionals who contributed to setting up and delivering the study at all of the 65 NHS trusts/Health boards and 25 research institutions across the UK, as well as all the supporting staff at the NIHR Clinical Research Network, Health Research Authority, Research Ethics Committee, Department of Health and Social Care, Public Health Scotland, and Public Health England, and support from the ISARIC Coronavirus Clinical Characterisation Consortium. We thank Kate Holmes at the NIHR Office for Clinical Research Infrastructure (NOCRI) for her support in coordinating the charities group. The PHOSP-COVID industry framework was formed to provide advice and support in commercial discussions, and we thank the Association of the British Pharmaceutical Industry as well as Ivana Poparic and Peter Sargent at NOCRI for coordinating this. We are very grateful to all the charities that have provided insight to the study: Action Pulmonary Fibrosis, Alzheimer’s Research UK, Asthma UK/British Lung Foundation, British Heart Foundation, Diabetes UK, Cystic Fibrosis Trust, Kidney Research UK, MQ Mental Health, Muscular Dystrophy UK, Stroke Association Blood Cancer UK, McPin Foundations, and Versus Arthritis. We thank the NIHR Leicester Biomedical Research Centre patient and public involvement group and the Long Covid Support Group.

