## Supplementary material for "Device-assessed sleep and physical activity in individuals recovering from a hospital admission for COVID-19: a prospective, multicentre study"

### Supplementary methods

#### Accelerometer data processing

Data were downloaded using GENEActiv PC software V.3.2 and accelerometer files were processed with R-package GGIR version 2.2-0 (<http://cran.r-project.org>).^1^

Signal processing in GGIR includes autocalibration using local gravity as a reference; detection of non-wear; calculation of the average magnitude of dynamic acceleration corrected for gravity (Euclidean Norm minus 1 *g* with negative values rounded up to zero, ENMO), averaged over 5-s epochs and expressed in milli-gravitational units (m*g*). Non-wear was imputed using the default setting, that is, invalid data were imputed by the average at similar time-points on different days of the week. Participants were excluded if post-calibration error was >0.01 *g* (10 m*g*), they had < 3 days of valid wear (defined as ≥16 h per day), or if wear data were not present for each 15-minute period of the 24-h cycle.

Sleep characteristics were obtained using a validated automated sleep detection algorithm.^2^ This algorithm facilitates detection of the sleep period time window (SPT-window) without the use of sleep diaries. The SPT-window refers to the time window starting at sleep onset and ending when waking up after the last sleep episode of the night.

Individual nights were excluded if the sleep window was <3 h, >12 h, or the number of sleep episodes per night was ≤5 or ≥30 as in previous research.^3^ In addition, nights with sleep efficiency <50% were visually inspected and unrealistic nights removed (e.g., sleep window clearly erroneously detected). Out of 13 348 nights, 1 452 (~11%) were excluded based on these criteria.

#### Definitions of physical activity and sleep variables

| ***Daily physical activity variables*** | **Definition** |
| --- | --- |
| Average acceleration (m*g)* | A proxy for physical activity volume |
| Moderate-to-vigorous intensity physical activity (minutes) | Accumulated in ≥1-minute bouts (>100 m*g*)^4^ |
| Light-intensity activity (minutes) | Time accumulated with an acceleration between 40 and 100 m*g*^5^ |
| Inactive time (minutes) | Time accumulated during the waking day below 40 m*g*^5^ |
| Intensity of the most active continuous 30 and 10 minutes/day (m*g*) | Acceleration value corresponding to the 25th percentile of the distribution during the most active period, therefore 75% of the time within these bouts is spent above this value |
| ***Weekly physical activity characteristics*** |  |
| Number of days per week with 10- and 30-minute continuous MVPA | Weekly variables were limited to participants with at least seven valid days of data. A continuous 10 (or 30) minute moderate-to-vigorous activity session is evident when the intensity corresponding with the 25th percentile of the distribution for the 10 (or 30) continuous minutes is >100 m*g* (e.g., walking), i.e., there is an allowance for up to 25% of time to drop below the threshold consistent with standard bout definitions (e.g., Troiano et al., 2008).^6^ |
| ***Sleep variables*** |  |
| Sleep duration (hours) | Total accumulated sleep within the sleep window |
| Sleep efficiency (%) | The ratio of sleep duration to the duration of the sleep window |
| Mid-sleep variability (SD in minutes) | Within-person standard deviation of mid-sleep time (mid-point between sleep onset and waking time). Sleep mid-point variability describes how variable people are in the timing of their sleep window. |

#### Disease definitions

WHO clinical progression scale,^7^ defined as:

i) Class 3-4 = no continuous supplemental oxygen needed,
ii) Class 5 = continuous supplemental oxygen only,
iii) Class 6 = continuous positive airway pressure ventilation (CPAP), bi-level positive airway pressure (BIPAP) or high flow nasal oxygen,
iv) Class 7-9 = invasive mechanical ventilation (IMV) or extra-corporeal membrane oxygenation (ECMO).

Four recovery clusters from a battery of patient reported outcomes and physical tests:^8^

1) Very severe mental and physical health impairment,
2) Severe mental and physical health impairment,
3) Moderate mental and physical health impairment with pronounced cognitive impairment,
4) Mild mental and physical health impairment

The battery of patient reported outcomes and physical tests used to create these clusters, which were assessed using validated tools, included:

| **Symptom** | **Method** |
| --- | --- |
| Breathlessness | Dyspnoea-12^9^ |
| Fatigue | FACIT (Functional Assessment of Chronic Illness Therapy)^10^ |
| Anxiety | GAD-7 (General Anxiety Disorder 7 Questionnaire)^11^ |
| Depression | PHQ-9 (Patient Health Questionnaire – 9)^12^ |
| Post-traumatic stress disorder | PCL-5 (Post Traumatic Stress Disorder Checklist)^13^ |
| Physical performance | SPPB (Short Physical Performance Battery)^14^ |
| Cognition | MoCA (Montreal Cognitive Assessment)^15^ |

#### Covariates

| **Covariate** | **Method** |
| --- | --- |
| Ethnicity | Ethnicity was recorded according to census definitions and classified as White (English, Welsh, Scottish, Northern Irish or British, Irish, Gypsy or Irish Traveller any other White background), Black (African, Caribbean, any other Black, African or Caribbean background), South Asian (Indian, Pakistani, Bangladeshi) or other (including mixed) ethnicities. |
| Comorbidity | Derived from a comprehensive medical history taken during the hospital admission and categorised as none, one or two or more comorbidities. |
| Index of Multiple Deprivation | Based on seven domains of deprivation: income, employment, education, skills and training, health and disability, crime, barriers to housing services, living environment and centred on participants postcodes and categorised in quintiles based on nationally derived thresholds. |

#### Comparative cohorts

The SMART Work and Life (SWL) study recruited adult office workers aged ≥18 years within local Councils in the Leicester, Manchester and Liverpool areas between 2018 and 2019. Details of the study have been published.^16^ Participants wore an Axivity AX3 (Axivity Ltd, Newcastle, UK) accelerometer their non-dominant wrist 24 h/day for 7 days, with data recorded at 100 Hz. Previous studies showed that physical activity and sleep outcomes from the GENEActiv and Axivity accelerometers are comparable when generated using methods as described above.^17,18^ Because the SWL cohort was younger, to match with the PHOSP sample, age was split into tertiles and only participants in the upper tertile were included in the analysis (***Supplementary Table S5***).

The CODEC study (Chronotype of Patients with Type 2 Diabetes and Effect on Glycaemic Control) recruited people with type 2 diabetes throughout the East Midlands between 2017 and 2019. The CODEC cohort study has been described in detail previously.^19,20^ Individuals wore the same accelerometer as in PHOSP-COVID (GENEActiv) for 7 days, with data recorded at 100 Hz. Characteristics of the CODEC sample, including the number included, were similar to the PHOSP sample (***Supplementary Table S5***).

### Supplementary results

#### Supplementary tables

**Supplementary Table S1: Participant characteristics by missing data status.**

|  |  | **Missing** | | **Complete** | |
| --- | --- | --- | --- | --- | --- |
| Variable |  | **Count** | **Column %** | **Count** | **Column %** |
| WHO disease severity class* | Class 3-4 | 87 | 24.0% | 139 | 19.4% |
|  | Class 5 | 123 | 34.0% | 255 | 35.7% |
|  | Class 6 | 69 | 19.1% | 116 | 16.2% |
|  | Class 7-9 | 83 | 22.9% | 205 | 28.7% |
| Recovery cluster | Cluster 1: Very severe | 45 | 12.4% | 86 | 12.0% |
|  | Cluster 2: Severe | 51 | 14.1% | 108 | 15.1% |
|  | Cluster 3: Moderate | 38 | 10.5% | 89 | 12.4% |
|  | Cluster 4: Mild | 112 | 30.9% | 238 | 33.3% |
|  | NA | 116 | 32.0% | 194 | 27.1% |
| Sex | Female | 131 | 36.2% | 253 | 35.4% |
|  | Male | 231 | 63.8% | 462 | 64.6% |
| Ethnicity | White | 211 | 58.3% | 499 | 69.8% |
|  | South Asian | 74 | 20.4% | 92 | 12.9% |
|  | Black | 40 | 11.0% | 49 | 6.9% |
|  | Mixed | 8 | 2.2% | 15 | 2.1% |
|  | Other | 19 | 5.2% | 28 | 3.9% |
|  | Missing | 10 | 2.8% | 32 | 4.5% |
| IMD Quintile** | 1 - most deprived | 80 | 22.1% | 135 | 18.9% |
|  | 2 | 76 | 21.0% | 166 | 23.2% |
|  | 3 | 75 | 20.7% | 127 | 17.8% |
|  | 4 | 55 | 15.2% | 141 | 19.7% |
|  | 5 - least deprived | 53 | 14.6% | 145 | 20.3% |
|  | Missing | 23 | 6.4% | 1 | 0.1% |
| **Continuous Variables** | | Mean | SD | Mean | SD |
| Age (years) | | 55 | 13 | 59 | 13 |
| BMI*** | | 32.1 | 7.5 | 31.0 | 6.8 |

*WHO clinical progression scale: 3-4 = no continuous supplemental oxygen needed, 5 = continuous supplemental oxygen only, 6 = continuous positive airway pressure ventilation (CPAP), bi-level positive airway pressure (BIPAP) or high flow nasal oxygen, 7-9 = invasive mechanical ventilation (IMV) or extra-corporeal membrane oxygenation (ECMO). ** IMD = Index of Multiple Deprivation; ***BMI = body mass index

**Supplementary Table S2. Sleep variables across WHO classes of acute disease severity and four recovery clusters excluding healthcare workers (N = 518).**

| **WHO classes** | | | | | | | | | | |
| --- | --- | --- | --- | --- | --- | --- | --- | --- | --- | --- |
|  | **Women** | | | | **Men** | | | |  |  |
| **Sleep variables** | **Class 3-4** | **Class 5** | **Class 6** | **Class 7-9** | **Class 3-4** | **Class 5** | **Class 6** | **Class 7-9** | **P for class** | **P for class x sex** |
| Sleep duration (hrs/night) | 6.94 (6.55, 7.33) | 7.00 (6.65, 7.36) | 7.01 (6.59, 7.60) | 6.78 (6.32, 7.24) | 6.64 (6.27, 7.01) | 6.68 (6.38, 6.98) | 6.67 (6.30, 7.04) | 6.63 (6.31, 6.94) | 0.723 | 0.878 |
| Sleep efficiency (%) | 87.07 (85.13, 89.01) | 87.09 (85.33, 88.86) | 86.56 (84.05, 89.06) | 86.19 (83.89, 88.48) | 86.63 (84.79, 88.48) | 85.81 (84.32, 87.30) | 85.96 (84.10, 87.82) | 86.26 (84.69, 87.83) | 0.854 | 0.799 |
| Sleep midpoint variability (minutes) | 91.37 (62.14, 120.59) | 87.35 (60.77, 113.92) | 74.72 (37.04, 112.39) | 115.81 (81.21, 150.42) | 100.59 (72.76, 128.42) | 86.56 (64.08, 109.04) | 107.12 (79.10, 135.13) | 85.73 (62.09, 109.38) | 0.616 | 0.103 |
| **Recovery clusters** | | | | | | | | | | |
|  | **Women** | | | | **Men** | | | |  |  |
| **Sleep variables** | **Cluster 4: Mild** | **Cluster 3: Moderate** | **Cluster 2: Severe** | **Cluster 1: Very Severe** | **Cluster 4: Mild** | **Cluster 3: Moderate** | **Cluster 2: Severe** | **Cluster 1: Very Severe** | **P for class** | **P for class x sex** |
| Sleep duration (hrs/night) | 7.18 (6.77, 7.58) | 6.92 (6.41, 7.43) | 7.14 (6.68, 7.61) | 6.70 (6.18, 7.22) | 6.70 (6.38, 7.01) | 6.52 (6.14, 6.91) | 7.06 (6.65, 7.47) | 6.76 (6.32, 7.20) | 0.144 | 0.356 |
| Sleep efficiency (%) | 86.85 (84.81, 88.88) | 87.07 (84.50, 89.63) | 87.20 (84.85, 89.54) | 86.34 (83.73, 88, 97) | 86.18 (84.59, 87.77) | 85.33 (83.39, 87.27) | 86.04 (84.32, 88.48) | 85.43 (83.19, 87.66) | 0.808 | 0.934 |
| Sleep midpoint variability (minutes) | 87.41 (56.94, 117.88) | 88.86 (50.31, 127.40) | 78.25 (43.05, 113.45) | 100.03 (60.72, 139.34) | 85.87 (62.05, 109.68) | 98.39 (69.31, 127.47) | 74.10 (42.94, 105.25) | 116.65 (83.13, 150.16) | 0.172 | 0.874 |

Data reported as marginal mean (95% CI). Adjusted for age, sex, ethnicity, deprivation, number of comorbidities, season of data collection, and number of wear nights (sleep variables). Models included 518 participants with complete cluster and covariate data.

**Supplementary Table S3: Associations of independent cluster variables with physical activity and sleep variables.**

| **Exposure** | **Outcome** | b (95% CI) | P for main effect | P for interaction by sex |
| --- | --- | --- | --- | --- |
| **Breathlessness (Dyspnoea-12)** | **Physical activity variables** |  |  |  |
|  | Physical activity volume (m*g*) | -0.75 (-1.25, -0.25) | 0.003 | 0.867 |
|  | Moderate to vigorous intensity physical activity (minutes/day) | -2.64 (-4.08, -1.20) | 0.001 | 0.351 |
|  | Light intensity physical activity (minutes/day) | -6.03 (-9.93, -2.14) | 0.002 | 0.380 |
|  | Inactivity (minutes/day) | 4.41 (-2.95, 11.77) | 0.240 | 0.653 |
|  | Intensity of the most active continuous 30 minutes (m*g*) | -4.37 (-7.6, -1.14) | 0.008 | 0.683 |
|  | Intensity of the most active continuous 10 minutes (m*g*) | -7.20 (-11.61, -2.79) | 0.001 | 0.573 |
|  | **Sleep variables** |  |  |  |
|  | Sleep duration (hrs/night) | 0.04 (-0.05, 0.12) | 0.405 | 0.585 |
|  | Sleep efficiency (%) | -0.23 (-0.65, 0.19) | 0.283 | 0.662 |
|  | Sleep midpoint variability (minutes) | 8.52 (2.12, 14.93) | 0.009 | 0.888 |
| **Fatigue (FACIT)** | **Physical activity variables** |  |  |  |
|  | Physical activity volume (m*g*) | -1.26 (-1.79, -0.72) | 0.001 | 0.762 |
|  | Moderate to vigorous intensity physical activity (minutes/day) | -4.15 (-5.67, -2.64) | 0.001 | 0.112 |
|  | Light intensity physical activity (minutes/day) | -8.94 (-13.07, -4.81) | 0.001 | 0.763 |
|  | Inactivity (minutes/day) | 10.47 (2.61, 18.33) | 0.009 | 0.750 |
|  | Intensity of the most active continuous 30 minutes (m*g*) | -7.29 (-10.63, -3.94) | 0.001 | 0.175 |
|  | Intensity of the most active continuous 10 minutes (m*g*) | -11.32 (-15.88, -6.75) | 0.001 | 0.099 |
|  | **Sleep variables** |  |  |  |
|  | Sleep duration (hrs/night) | 0.02 (-0.06, 0.11) | 0.594 | 0.661 |
|  | Sleep efficiency (%) | -0.05 (-0.50, 0.40) | 0.834 | 0.805 |
|  | Sleep midpoint variability (minutes) | 11.58 (4.75, 18.41) | 0.001 | 0.409 |

Data reported as beta-coefficients (95% CI). Adjusted for age, sex, ethnicity, deprivation, number of comorbidities, season of data collection, and number of wear days (physical activity variables) or wear nights (sleep variables). FACIT = Functional Assessment of Chronic Illness Therapy.

| **Exposure** | **Outcome** | b (95% CI) | P for main effect | P for interaction by sex |
| --- | --- | --- | --- | --- |
| **Physical performance (SPPB)** | **Physical activity variables** |  |  |  |
|  | Physical activity volume (m*g*) | 0.90 (0.34, 1.47) | 0.002 | 0.058 |
|  | Moderate to vigorous intensity physical activity (minutes/day) | 3.65 (2.00, 5.29) | 0.001 | 0.037 |
|  | Light intensity physical activity (minutes/day) | 5.59 (1.33, 9.85) | 0.010 | 0.164 |
|  | Inactivity (minutes/day) | -15.06 (-23.17, -6.95) | 0.001 | 0.426 |
|  | Intensity of the most active continuous 30 minutes (m*g*) | 7.21 (3.67, 10.76) | 0.001 | 0.016 |
|  | Intensity of the most active continuous 10 minutes (m*g*) | 9.52 (4.64, 14.40) | 0.001 | 0.034 |
|  | **Sleep variables** |  |  |  |
|  | Sleep duration (hrs/night) | 0.01 (-0.09, 0.09) | 0.970 | 0.708 |
|  | Sleep efficiency (%) | -0.12 (-0.59, 0.34) | 0.604 | 0.530 |
|  | Sleep midpoint variability (minutes) | -2.74 (-9.94, 4.47) | 0.457 | 0.055 |
| **Cognition (MoCA)** | **Physical activity variables** |  |  |  |
|  | Physical activity volume (m*g*) | -0.03 (-0.60, 0.54) | 0.922 | 0.554 |
|  | Moderate to vigorous intensity physical activity (minutes/day) | 0.98 (-0.72, 2.68) | 0.258 | 0.983 |
|  | Light intensity physical activity (minutes/day) | -2.52 (-6.84, 1.80) | 0.253 | 0.600 |
|  | Inactivity (minutes/day) | 3.08 (-5.19, 11.35) | 0.466 | 0.953 |
|  | Intensity of the most active continuous 30 minutes (m*g*) | 2.12 (-1.68, 5.91) | 0.274 | 0.637 |
|  | Intensity of the most active continuous 10 minutes (m*g*) | 4.28 (-0.92, 9.48) | 0.107 | 0.742 |
|  | **Sleep variables** |  |  |  |
|  | Sleep duration (hrs/night) | 0.06 (-0.03, 0.16) | 0.180 | 0.681 |
|  | Sleep efficiency (%) | 0.40 (-0.07, 0.87) | 0.092 | 0.371 |
|  | Sleep midpoint variability (minutes) | -6.92 (-14.13, 0.29) | 0.060 | 0.467 |

Data reported as beta-coefficients (95% CI). Adjusted for age, sex, ethnicity, deprivation, number of comorbidities, season of data collection, and number of wear days (physical activity variables) or wear nights (sleep variables). SPPB = short physical performance battery, MoCA = Montreal Cognitive Assessment.

| **Exposure** | **Outcome** | b (95% CI) | P for main effect | P for interaction by sex |
| --- | --- | --- | --- | --- |
| **Anxiety (GAD-7)** | **Physical activity variables** |  |  |  |
|  | Physical activity volume (m*g*) | -0.30 (-0.82, 0.22) | 0.256 | 0.114 |
|  | Moderate to vigorous intensity physical activity (minutes/day) | -1.57 (-3.06, -0.09) | 0.038 | 0.832 |
|  | Light intensity physical activity (minutes/day) | -3.52 (-7.56, 0.52) | 0.088 | 0.314 |
|  | Inactivity (minutes/day) | 0.19 (-7.46, 7.84) | 0.961 | 0.900 |
|  | Intensity of the most active continuous 30 minutes (m*g*) | -2.56 (-5.84, 0.72) | 0.126 | 0.804 |
|  | Intensity of the most active continuous 10 minutes (m*g*) | -4.35 (-8.82, 0.11) | 0.056 | 0.772 |
|  | **Sleep variables** |  |  |  |
|  | Sleep duration (hrs/night) | 0.02 (-0.07, 0.11) | 0.645 | 0.854 |
|  | Sleep efficiency (%) | -0.37 (-0.81, 0.06) | 0.093 | 0.984 |
|  | Sleep midpoint variability (minutes) | 9.45 (2.76, 16.15) | 0.006 | 0.327 |
| **Depression (PHQ-9)** | **Physical activity variables** |  |  |  |
|  | Physical activity volume (m*g*) | -0.58 (-1.11, -0.05) | 0.031 | 0.341 |
|  | Moderate to vigorous intensity physical activity (minutes/day) | -2.35 (-3.86, -0.85) | 0.002 | 0.318 |
|  | Light intensity physical activity (minutes/day) | -5.91 (-9.99, -1.82) | 0.005 | 0.369 |
|  | Inactivity (minutes/day) | 4.34 (-3.41, 12.10) | 0.272 | 0.901 |
|  | Intensity of the most active continuous 30 minutes (m*g*) | -4.21 (-7.54, -0.88) | 0.013 | 0.791 |
|  | Intensity of the most active continuous 10 minutes (m*g*) | -7.01 (-11.55, -2.47) | 0.002 | 0.606 |
|  | **Sleep variables** |  |  |  |
|  | Sleep duration (hrs/night) | -0.02 (-0.10, 0.07) | 0.732 | 0.419 |
|  | Sleep efficiency (%) | -0.46 (-0.90, -0.02) | 0.039 | 0.774 |
|  | Sleep midpoint variability (minutes) | 10.35 (3.55, 17.15) | 0.003 | 0.762 |

Data reported as beta-coefficients (95% CI). Adjusted for age, sex, ethnicity, deprivation, number of comorbidities, season of data collection, and number of wear days (physical activity variables) or wear nights (sleep variables). GAD-7 = General Anxiety Disorder 7 Questionnaire, PHQ-9 = Patient Health Questionnaire – 9.

| **Exposure** | **Outcome** | b (95% CI) | P for main effect | P for interaction by sex |
| --- | --- | --- | --- | --- |
| **Post-traumatic stress disorder (PCL-5)** | **Physical activity variables** |  |  |  |
|  | Physical activity volume (m*g*) | -0.66 (-1.16, -0.15) | 0.011 | 0.575 |
|  | Moderate to vigorous intensity physical activity (minutes/day) | -2.53 (-3.98, -1.07) | 0.001 | 0.226 |
|  | Light intensity physical activity (minutes/day) | -5.35 (-9.28, -1.43) | 0.008 | 0.551 |
|  | Inactivity (minutes/day) | 3.68 (-3.8, 11.15) | 0.335 | 0.656 |
|  | Intensity of the most active continuous 30 minutes (m*g*) | -4.06 (-7.29, -0.83) | 0.014 | 0.768 |
|  | Intensity of the most active continuous 10 minutes (m*g*) | -6.56 (-10.98, -2.15) | 0.004 | 0.529 |
|  | **Sleep variables** |  |  |  |
|  | Sleep duration (hrs/night) | -0.01 (-0.1, 0.07) | 0.786 | 0.286 |
|  | Sleep efficiency (%) | -0.26 (-0.68, 0.17) | 0.242 | 0.387 |
|  | Sleep midpoint variability (minutes) | 12.57 (6.02, 19.11) | 0.001 | 0.237 |

Data reported as beta-coefficients (95% CI). Adjusted for age, sex, ethnicity, deprivation, number of comorbidities, season of data collection, and number of wear days (physical activity variables) or wear nights (sleep variables). PCL-5 = Post Traumatic Stress Disorder Checklist.

**Supplementary Table S4:** **Associations of physical performance with moderate to vigorous intensity physical activity and intensity of the most active continuous 30/10 minutes stratified by sex.**

|  |  | **Women** | | **Men** | |
| --- | --- | --- | --- | --- | --- |
| **Exposure** | **Outcome** | b (95% CI) | p for main effect | b (95% CI) | p for main effect |
| **Physical performance (SPPB)** |  |  |  |  |  |
|  | Moderate to vigorous intensity physical activity (minutes/day) | 2.11 (0.07, 4.14) | 0.042 | 4.51 (2.22, 6.81) | 0.001 |
|  | Intensity of the most active continuous 30 minutes (m*g*) | 2.92 (-0.65, 6.48) | 0.109 | 9.90 (4.79, 15.00) | 0.001 |
|  | Intensity of the most active continuous 10 minutes (m*g*) | 5.26 (-0.41, 10.92) | 0.069 | 12.04 (5.19, 18.90) | 0.001 |

Data reported as beta-coefficients (95% CI). Adjusted for age, ethnicity, deprivation, number of comorbidities, season of data collection, and number of wear days. SPPB = short physical performance battery.

**Supplementary Table S5: Participant characteristics in the CODEC and SWL compared to the PHOSP-COVID cohorts.**

|  |  | **PHOSP** | | **CODEC** | | **SWL** | |
| --- | --- | --- | --- | --- | --- | --- | --- |
| **Categorical variables** |  | **Count** | **Column %** | **Count** | **Column %** | **Count** | **Column%** |
| Sex | Female | 253 | 35.4% | 236 | 34.5% | 165 | 71.1% |
|  | Male | 462 | 64.6% | 449 | 65.5% | 67 | 28.9% |
| Ethnicity | White | 499 | 69.8% | 576 | 84.1% | 175 | 75.4% |
|  | South Asian | 92 | 12.9% | 80 | 11.7% | 44 | 19.0% |
|  | Black | 49 | 6.9% | 21 | 3.1% | 5 | 2.2% |
|  | Other | 43 | 6.0% | 8 | 1.2% | 8 | 3.4% |
|  | Missing | 32 | 4.5% | 0 | 0.0% | 0 | 0.0% |
| IMD (quintile)* | 1 | 135 | 18.9% | 95 | 13.9% | 25 | 10.8% |
|  | 2 | 166 | 23.2% | 96 | 14.0% | 33 | 14.2% |
|  | 3 | 127 | 17.8% | 110 | 16.1% | 47 | 20.3% |
|  | 4 | 141 | 19.7% | 156 | 22.8% | 61 | 26.3% |
|  | 5 | 145 | 20.3% | 224 | 32.7% | 64 | 27.6% |
|  | Missing | 1 | 0.1% | 4 | 0.6% | 2 | 0.9% |
| Comorbidities | No comorbidity | 205 | 28.7% | 95 | 13.9% | 178 | 76.7% |
|  | 1 comorbidity | 145 | 20.3% | 186 | 27.2% | 47 | 20.3% |
|  | 2+ comorbidities | 365 | 51.0% | 404 | 59.0% | 7 | 3.0% |
| Continuous variables | | Mean | SD | Mean | SD | Mean | SD |
| Age (years) | | 59 | 13 | 64 | 8 | 55.8 | 3.4 |
| BMI (kg/m^2^)** | | 31 | 6.8 | 30.9 | 5 | 26.9 | 5.5 |

* IMD = Index of Multiple Deprivation; **BMI = body mass index

**Supplementary Table S6: Physical activity and sleep variables in the CODEC and SWL compared to the PHOSP-COVID cohorts.**

|  | **PHOSP** | | **CODEC** | | **SWL** | | **P for group** | **P for group x sex** |
| --- | --- | --- | --- | --- | --- | --- | --- | --- |
|  | **Women** | **Men** | **Women** | **Men** | **Women** | **Men** |  |  |
| **Physical activity variables** |  |  |  |  |  |  |  |  |
| Physical activity volume (m*g*) | 20.9  (19.5, 22.2) | 22.0  (20.8, 23.2) | 22.2  (20.9, 23.5) | 22.9  (21.8, 24.1) | 23.3  (22.0, 24.7) | 26.0  (24.2, 27.8) | <0.001 | 0.218 |
| Moderate to vigorous intensity physical activity (minutes/day) | 17.7  (13.3, 22.0) | 25.9  (22.0, 29.9) | 19.7  (15.6, 23.9) | 26.1  (22.4, 29.9) | 30.9  (26.5, 35.3) | 44.7  (38.7, 50.7) | <0.001 | 0.118 |
| Light intensity physical activity (minutes/day) | 168.5  (157.8, 179.2) | 158.7  (148.8, 168.5) | 184.3  (174.3, 194.3) | 177.0  (167.8, 186.1) | 176.0  (165.5, 186.5) | 156.2  (141.9, 170.6) | 0.004 | 0.340 |
| Inactivity (minutes/day) | 738.8  (717.9, 759.6) | 765.5  (746.2, 784.8) | 733.4  (713.8, 753.0) | 730.7  (712.8, 748.6) | 740.7  (720.2, 761.2) | 768.0  (740.0, 796.0) | 0.051 | 0.020 |
| Intensity of the most active continuous 30 minutes (mg) | 37.7  (27.6, 47.8) | 48.3  (39.1, 57.5) | 46.8  (37.3, 56.4) | 51.8  (43.2, 60.4) | 52.1  (42.0, 62.2) | 103.4  (89.6, 117.2) | <0.001 | <0.001 |
| Intensity of the most active continuous 10 minutes (m*g*) | 63.3  (50.1, 76.5) | 79.5  (67.4, 91.6) | 68.9  (56.4, 81.5) | 77.7  (66.4, 89.1) | 88.0  (74.8, 101.3) | 181.8  (163.7, 200.0) | <0.001 | <0.001 |
| **Sleep variables** |  |  |  |  |  |  |  |  |
| Sleep duration (hr/night) | 6.74  (6.52, 6.96) | 6.38  (6.17, 6.59) | 6.57  (6.37, 6.78) | 6.46  (6.27, 6.65) | 6.48  (6.25, 6.70) | 6.14  (5.84, 6.44) | 0.092 | 0.114 |
| Sleep efficiency (%) | 83.5  (82.3, 84.7) | 82.7  (81.6, 83.9) | 88.4  (87.3, 89.5) | 86.5  (85.5, 87.5) | 88.8  (87.6, 90.0) | 87.4  (85.6, 89.0) | <0.001 | 0.231 |
| Sleep midpoint variability (minutes) | 138.2  (124.7, 151.7) | 135.7  (122.9, 148.4) | 37.1  (24.4, 49.9) | 31.6  (20.0, 43.1) | 36.2  (22.7, 49.7) | 46.7  (28.3, 65.0) | <0.001 | 0.333 |

Data reported as marginal mean (95% CI). Adjusted for age, sex, ethnicity, deprivation, number of comorbidities, season of data collection, and number of wear days (physical activity variables) or wear nights (sleep variables).

M10 sessions p for group <0.001, group*sex 0.074

M30 sessions p for group < 0.001 group*sex 0.069

#### Supplementary figures

**Supplementary Figure S1: Flow of data inclusion**

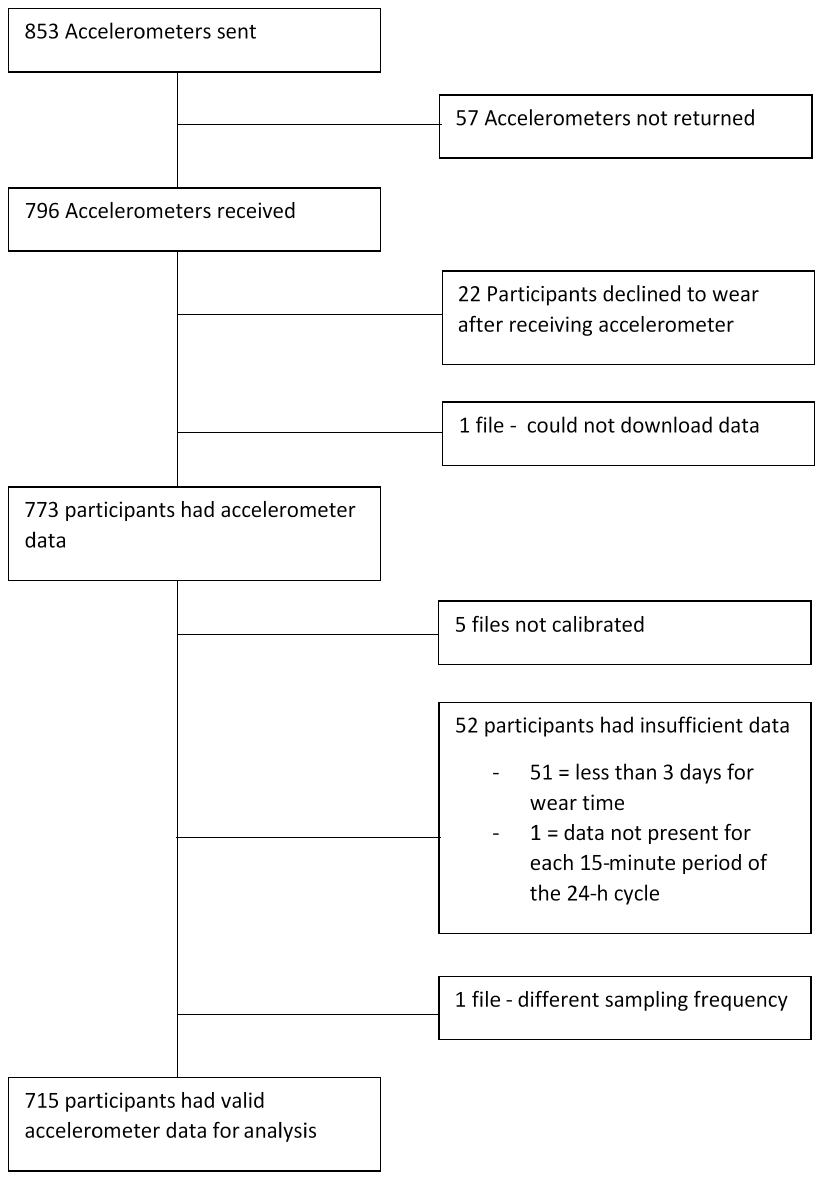

**Supplementary Figure S2: Odds ratios of not meeting 150 minutes of MVPA per week across acute illness severity.**

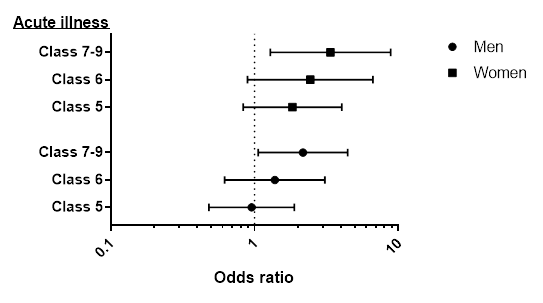

Data reported as odds ratios (95% CI). Reference group: Class 3-4. Analysis adjusted for age, sex, ethnicity, deprivation, number of comorbidities, season of data collection, and number of wear days. p for class =0.018, p for class x sex <0.001.

**Supplementary Figure S3: Odds ratios of not meeting 150 minutes of MVPA per week across recovery clusters.**

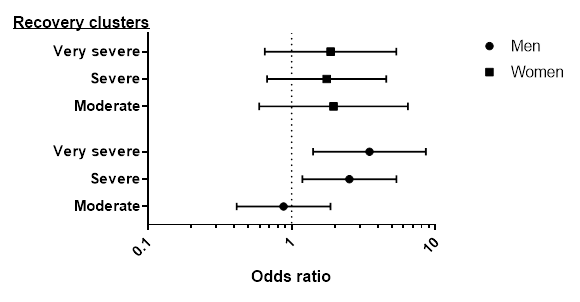

Data reported as odds ratios (95% CI). Reference group: Mild. Analysis adjusted for age, sex, ethnicity, deprivation, number of comorbidities, season of data collection, and number of wear days. p for class =0.012, p for cluster x sex =0.047.

Data display the proportion within each study achieving 0, 1, 2 and 3-7 days per week with a bout of 10 minutes (top panel) or 30 minutes (bottom) at least moderate-intensity physical activity. Analysis adjusted for age, sex, ethnicity, deprivation, number of comorbidities, season of data collection and number of wear days. 10-minute bouts: p for difference by studies = <0.001, p for difference by sex <0.001, p for sex x group = 0.074. 30-minute bouts: p for difference by studies = <0.001, p for difference by sex <0.001, p for sex x group = 0.069

**Supplementary Figure S4: proportion of participants within PHOSP, CODEC and SWL undertaking continuous bouts of physical activity.**

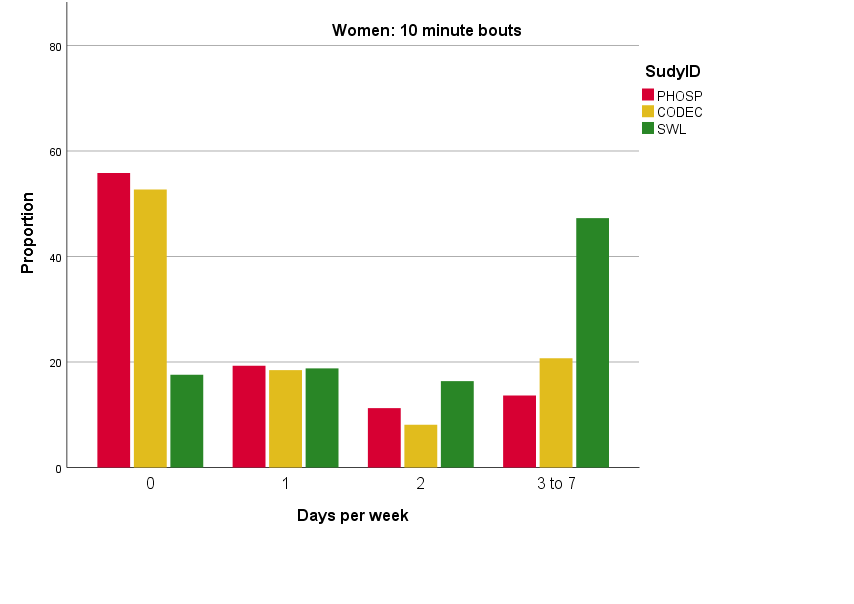

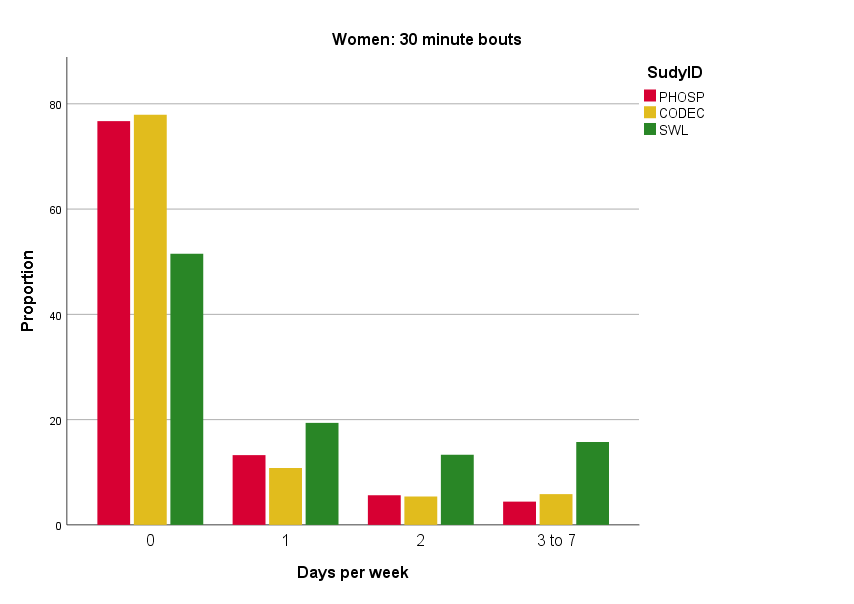

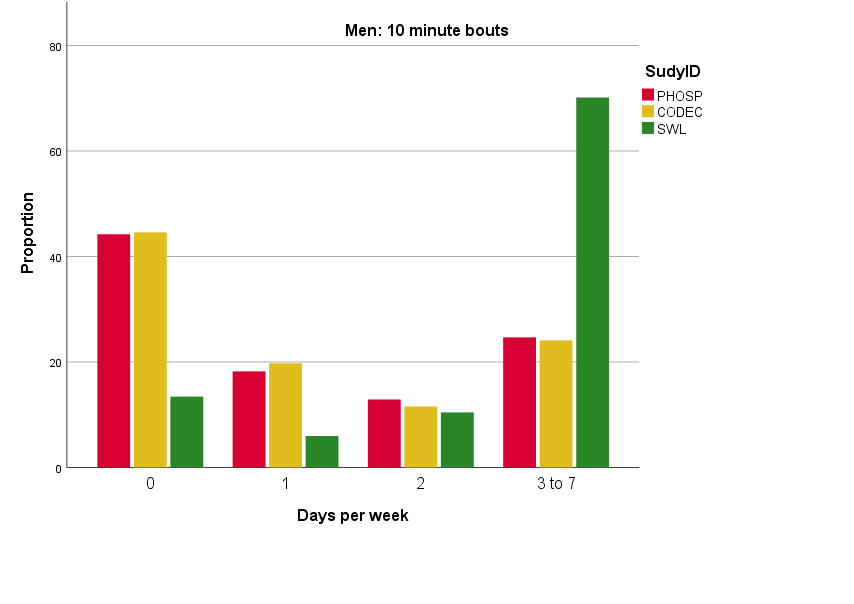

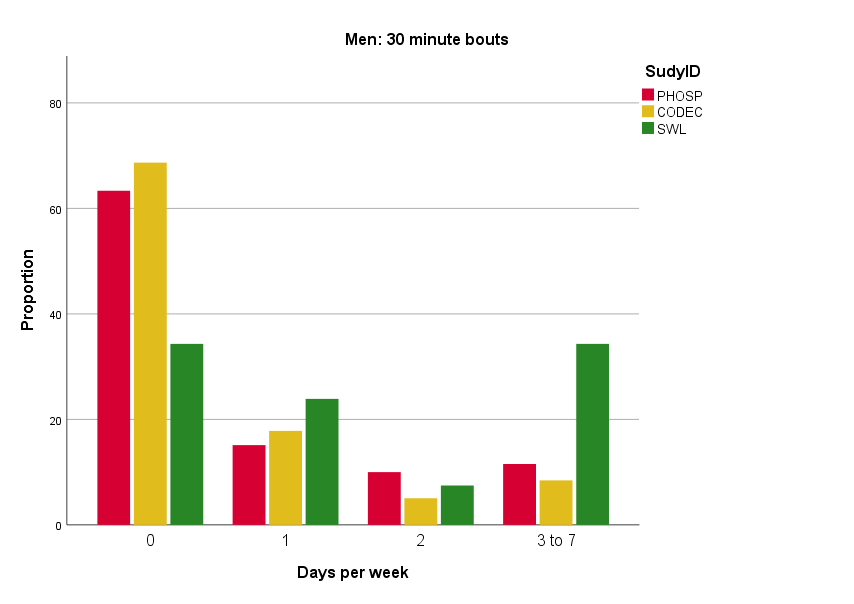
